## Supplementary Material for "Surveillance and control efficacy of the Bergerac, France, 2025 chikungunya outbreak"

Sandeep Tegar (https://orcid.org/0009-0003-2445-9860)[1,2*], Guillaume Lacour (https://orcid.org/0000-0002-9588-6064)[3], Antoine Mignotte (https://orcid.org/0000-0001-7433-1994)[3], Bethan V. Purse (https://orcid.org/0000-0001-5140-2710)[1], Christina A. Cobbold (https://orcid.org/0000-0001-8814-7688)[2], Dominic P. Brass (https://orcid.org/0000-0002-4900-9124)[1] & Steven M. White (https://orcid.org/0000-0002-3192-9969)[1*]

### Contents

1. Model Overview
2. Date of Primary Case Introduction
3. References

### S1. Model Overview

The model framework used to predict the chikungunya outbreak Bergerac, Dordogne, France 2025 is described by Brass and colleagues [1] for dengue virus. Here, the model is parameterised for chikungunya virus (CHIKV) transmission by incorporating the temperature-dependent relationships for *Ae. albopictus* and CHIKV vector competence and extrinsic incubation period and has been additionally validated against historic Italian CHIKV outbreaks [2] (Tegar et al. In Prep.).

To model the trait and population dynamics of Aedes albopictus, the model uses a continuous time delay differential equation approach [3] with an additional phenotypic structure to represent the effects of within and between generation plasticity [4]. This approach is flexible and able to incorporate the effect, both instantaneous and delayed, of environmental stressors on multiple traits, making it ideal for accounting for the complex life history of *Ae. albopictus*. Following Brass and colleagues [3], for a generic stage-structured population, $N$, the density of individuals in life stage $i$ and phenotypic class $j$ at time $t$ is described by a system of equations given by

$$\frac{dN_{i,j}(t)}{dt} = R_{i,j}(t) - M_{i,j}(t) - D_{i,j}(t),$$

for $i \in 1, \cdots, n$ and $j \in 1, \cdots, m$. Here, $R_{i,j}(t)$, $M_{i,j}(t)$ and $D_{i,j}(t)$ are functional forms describing the recruitment, maturation and death rate of individuals in life stage $i$ and phenotypic class $j$, respectively. These are often dependent on biotic and abiotic environments, parameterised by experimentally derived reaction norms describing the environment-trait relationship [1, 4].

The *Ae. albopictus* model predicts the dynamics of a population of mosquitoes arising from a single water body of fixed dimensions (Supplementary Figure S1). The model inputs are environmental variables (temperature, precipitation, evaporation and photoperiod) from the location being simulated and the outputs are predictions of population and trait dynamics (see main text). Adult mosquitoes oviposit eggs either onto the surface of the water or around the sides of the habitat, with the proportion of eggs being placed around the side of the habitat increasing as water level decreases. The eggs placed into or around the water body express either a diapausing ($E_D$) or non-diapausing ($E_\gamma$) phenotype which is determined by a maternal effect in response to falling temperatures and decreasing photoperiod. Once development is complete, both diapausing and non-diapausing eggs either become quiescent ($E_Q$) to survive dry periods or immediately hatch into larvae ($L$). Quiescence continues until the dormant egg is inundated by precipitation after which it immediately hatches. The development and survival of eggs are assumed to be temperature dependent, with diapausing eggs also being photoperiod dependent. The water body only varies in response to changes in temperature, the accumulation of precipitation, and through evaporation of standing water and is otherwise identical in every respect between locations.

Larval mosquitoes ($L$) compete for available resources in the aquatic habitat, consisting of a single larval class. Available resource is assumed to be consumed in its entirety and to be replenished daily, representing the product of temperature-dependent metabolic processes in the larval environment. Once larval development is complete pupation begins with the development of pupae (implicitly modelled due to the lack of density-dependence). The container habitats are vulnerable to flushing, a process whereby the body of water overflows and individuals are swept away. Flushing is modelled by increasing the mortality of larvae and pupae whenever the height of the water in the habitat exceeds the height of the container and rainfall is sufficiently intense. Further, the containers are also susceptible to drying out, and whenever all water within the container evaporates, all non-quiescent juveniles are assumed to die out.

Adult mosquitoes ($A$) experience developmental plasticity in response to their experience of temperature and intraspecific competition as larvae. Each individual's experience of the average temperature and the average food available per larvae per day over the course of the larval period is used to predict that individual's wing length. We discretise adults into a large number of coexisting sub-classes, with each adult class determined by their wing length ($A_i, i = 1, \cdots, m$), thus defining the adult phenotypic structure. Wing length is then used to determine the fecundity and longevity of that individual as an adult. Both fecundity and longevity are then further modified by the current temperature meaning that within the model adult traits respond to both current and historic environmental conditions. The production of cold resistant diapausing eggs is triggered when falling temperatures and photoperiod reach a critical threshold.

The model for the population and trait dynamics of *Ae. albopictus* is incorporated into a compartmental susceptible-exposed-infected-resistant (SEIR) model for dengue virus vectored by *Ae. albopictus* (Fig. S1). The human population is partitioned into those susceptible to infection ($H_S$), those infected ($H_I$), and those resistant to infection due to having recovered ($H_R$) – the exposed class is implicitly modelled. The size of the human population is constant, where the population density is estimated as described in the main text.

Mosquitoes are assumed to bite at a temperature-dependent rate that is inversely proportional to the length of the gonotrophic cycle. The proportion of uninfected mosquitoes of a given wing length ($A_i$) that become infected ($I_i$) after biting an infected human ($H_I$) is temperature dependent. After a temperature-dependent extrinsic incubation period, an infected mosquito can bite and transmit the chikungunya virus to an uninfected human ($H_S$). After the intrinsic incubation period, the infected human can transmit the infection to new mosquitoes and recovers from the infection after a fixed recovery period.

The specific form of delayed differential equations, along with the mosquito environmentally-dependent parameters are fully described by Brass and colleagues [1], and chikungunya virus parameters are available upon request [2] (Tegar et al. In Prep.).

Model code is available on GitHub (https://doi.org/10.5281/zenodo.17534800).

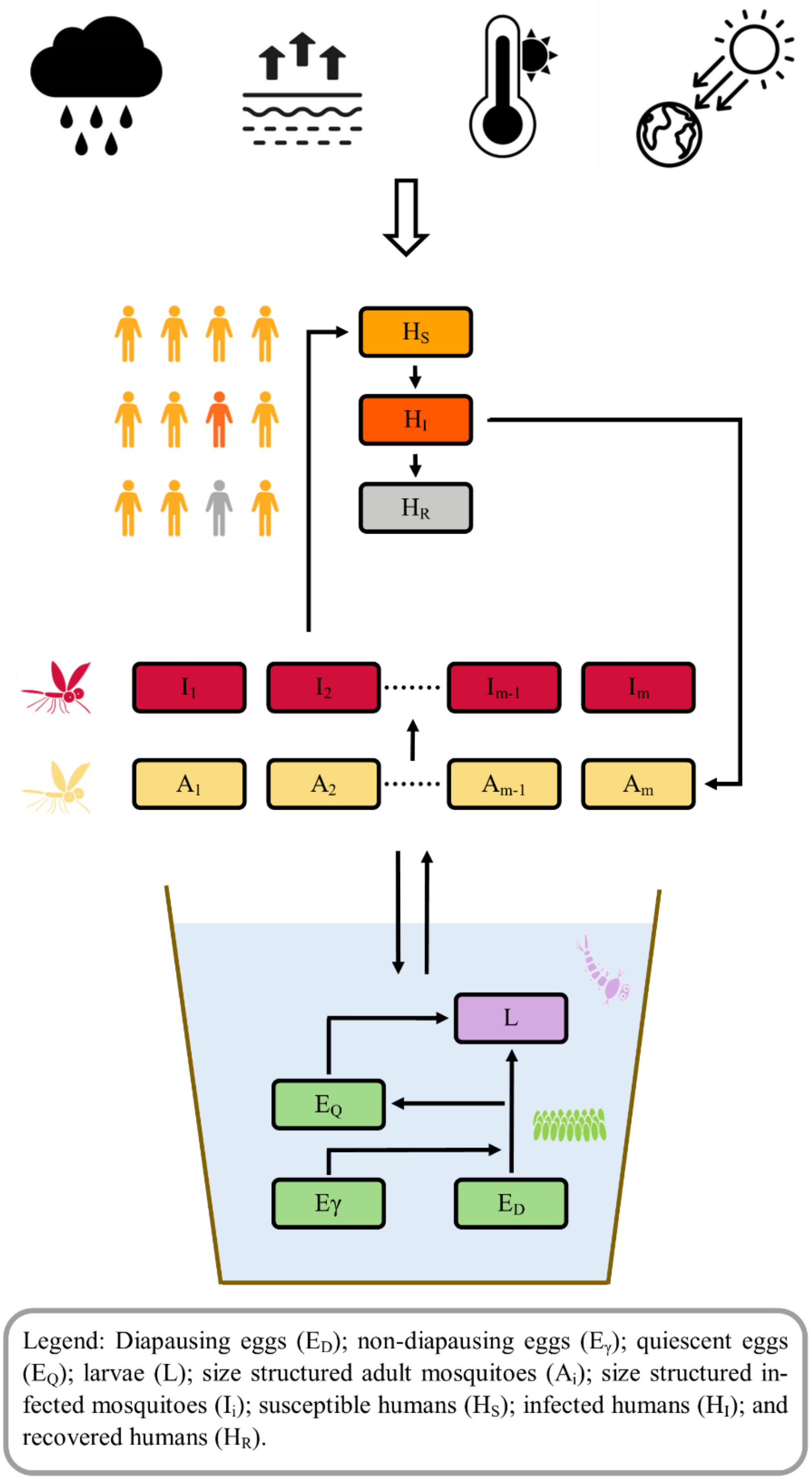


**Supplementary Figure S1:** A schematic of the stage and phenotypically structured delay differential equation epidemiological model [1]. The boxes and arrows indicate the flow between classes, the rates of which are determined by environmental variables (precipitation, temperature, evaporation and photoperiod). Image adapted from [5].

### S2. Date of Primary Case Introduction

We evaluated possible dates of primary case introduction in Bergerac ranging from 1 May 2025 to 30 June 2025. The optimal date was selected based on root mean square error (RMSE) minimisation. For each candidate date within this range, we also simulated the final outbreak size and assessed the variability in the resulting outbreak size (Supplementary Figure S2). The estimated date of primary case introduction corresponds well with the observed final outbreak size.

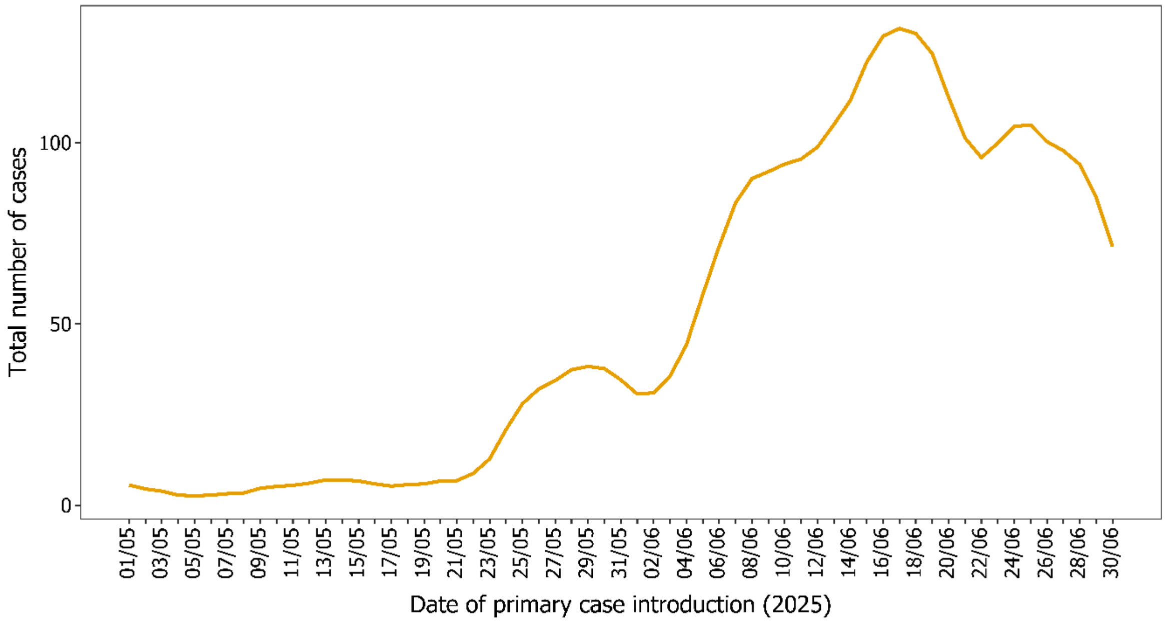


**Supplementary Figure S2:** Simulated final outbreak size for different dates of primary introduction of chikungunya infection in Bergerac, France, 2025.

### References


1. Brass, D.P., C.A. Cobbold, B.V. Purse, D.A. Ewing, A. Callaghan, and S.M. White, *Role of vector phenotypic plasticity in disease transmission as illustrated by the spread of dengue virus by Aedes albopictus.* Nature Communications, 2024. **15**(1): p. 7823.
2. Tegar, S., D.P. Brass, B.V. Purse, A. Mignotte, G. Lacour, C.A. Cobbold, and S.M. White, *Modelling and forecasting a potential chikungunya outbreak in France, 2025.* medRxiv, 2025: p. 2025.08.12.25333506.
3. Nisbet, R.M. and W.S.C. Gurney, *The systematic formulation of population models for insects with dynamically varying instar duration.* Theoretical Population Biology, 1983. **23**(1): p. 114-135.
4. Brass, D.P., C.A. Cobbold, D.A. Ewing, B.V. Purse, A. Callaghan, and S.M. White, *Phenotypic plasticity as a cause and consequence of population dynamics.* Ecology Letters, 2021. **24**(11): p. 2406-2417.
5. White, S.M., S. Tegar, B.V. Purse, C.A. Cobbold, and D.P. Brass, *Modelling the Lodi, 2023 and Fano 2024, Italy Dengue Outbreaks: The Effects of Control Strategies and Environmental Extremes.* Transboundary and Emerging Diseases, 2025. **2025**(1): p. 5542740.
